# Operative Time Heterogeneity in Laparoscopic Cholecystectomy at High Altitude: Surgeon Variability as a Modifiable Factor under Hypoxic Stress

**DOI:** 10.64898/2026.08.19.26360503

**Authors:** Guoliang Ren, Zhiqiang Wang, Zhongfeng Dang, Wei Su, Yabing Ma, Ping Li, Dongde Ji, Liansheng Li, Junlin Gao

**Affiliations:** Department of Hepatobiliary and Pancreatic Surgery, Qinghai Red Cross Hospital, Xining, Qinghai 810000, China; Ultrasound and Electrocardiography Center, Gansu Hospital of Sun Yat-sen University Cancer Center, Lanzhou, Gansu 730050, China

**Keywords:** Laparoscopic cholecystectomy, High altitude, Operative time, Surgeon variability, Surgical quality control

## Abstract

**Objective:** To quantify inter-surgeon heterogeneity in operative efficiency of laparoscopic cholecystectomy (LC) at high altitude and identify independent determinants of operative time.

**Methods:** A single-center retrospective cohort study at the participating hospital (2,260 m altitude) included 591 elective LC cases by 7 surgeons (2020–2023). One-way ANOVA, multivariate regression with log-transformed operative time, nested model comparison, and ICC quantified surgeon versus baseline factor contributions.

This study followed the STROBE (Strengthening the Reporting of Observational Studies in Epidemiology) reporting guidelines [16]; the checklist is provided in the supplementary materials.

**Results:** Inter-surgeon operative time differed significantly (F = 7.16, P < 0.001, η^2^ = 0.069), with 12.64 min (24.0%) gap between fastest and slowest surgeons. Regression (Adj R^2^ = 0.075, P < 0.001) identified age (β = 0.0029/yr, P = 0.009), male sex (β = 0.063, P = 0.019), and surgeon identity (Surgeon G β = 0.244, P < 0.001; Surgeon F β = 0.233, P = 0.001; Surgeon E β = 0.214, P = 0.018) as independent predictors. BMI showed a trend (P = 0.069). Nested comparison showed surgeon factors explained 2.02-fold more variance than all baseline factors combined (ΔR^2^ = 0.063 vs R^2^ = 0.031; ICC = 0.074). The F-statistic exceeded the F ≈ 2–4 range reported in plain-cohort LC studies.

**Conclusion:** Surgeon variability is the dominant modifiable determinant of operative time heterogeneity in high-altitude LC, contributing 2.02-fold more variance than patient baseline factors. Standardization of surgical technique represents a potentially important intervention target for efficiency improvement.

## 1. Introduction

Laparoscopic cholecystectomy (LC) is the standard minimally invasive procedure for symptomatic cholelithiasis[1,2], and operative time and perioperative resource consumption are core indicators of surgical care quality[3,4]. The determinants of LC efficiency and quality control frameworks in plain regions are well established, but in high-altitude environments (defined by WHO as >1,500 m), the low-pressure, hypoxic conditions may differentially affect patients’ tissue tolerance and surgeons’ operative error margins[5,6,7].

The impact of high-altitude hypoxia on LC procedures is not limited to a single physiological pathway but simultaneously disturbs tissue perfusion homeostasis, the coagulation–hemorrhage balance, and surgeons’ cognitive finesse[6,8,9], collectively compressing the operative tolerance space. Under such multidimensional homeostatic disruption, subtle inter-surgeon variations that can be absorbed by compensatory mechanisms at sea level may lose their compensatory reserve and manifest as quantifiable operative time differences[10].

Inter-surgeon operative time differences in plain cohorts are typically small, with F-statistics mostly in the 2–4 range[3,11,12] (direct comparison across studies is only directionally informative due to design heterogeneity). If the high-altitude hypoxic environment does amplify operative differences, an elevated F-statistic should be observable in plateau cohorts. This study leverages a consecutive cohort of 591 elective LC cases at high altitude (7 primary surgeons) to test this hypothesis in a falsifiable manner: if the plateau F-statistic does not exceed the plain range, the amplifier hypothesis is rejected. Nested model comparison is also used to quantify the relative contributions of surgeon versus baseline factors to operative time variance.

## 2. Materials and Methods

### 2.1 Study Design and Participants

Patients undergoing elective laparoscopic cholecystectomy (LC) between January 1, 2020 and December 31, 2023 at the Department of Hepatobiliary and Pancreatic Surgery, the participating hospital were enrolled. This center is located in Xining at approximately 2,260 m altitude, within the WHO-defined high-altitude physiological impact zone (>1,500 m).

#### Inclusion criteria

1. Age ≥18 years;
2. Complete electronic medical records, surgical records, anesthesia records, and HIS billing data;
3. Standard four-port or three-port elective LC (no conversion to open surgery, no concurrent biliary procedures).

#### Exclusion criteria

1. Conversion to open surgery;
2. Concomitant choledocholithiasis or biliary anomalies;
3. Hepatobiliary malignancy or severe hepatic/renal impairment;
4. Missing key research variables;
5. Low-volume surgeons (<7 cases) to ensure statistical stability of inter-group comparisons (potential selection bias is discussed in the Limitations).

Enrollment flow: 613 cases initially recorded → 5 cases excluded as outliers (unparseable dates or negative length of stay) → 608 valid cases after cleaning → top 7 surgeons by volume selected → final 591 cases. This study is based on the same clinical cohort as Series II (epidemiological characteristics analysis) and Series III (health economics analysis), but focuses on an independent scientific question: this study examines surgical efficiency heterogeneity across 7 surgeons, Series II investigates disease risk-factor remodeling in 605 patients, and Series III develops cost control models based on 605 patients with an extended time window. The sample size differences across studies (Series I: n=591, focusing on the top 7 surgeons by volume; Series II: n=605, all eligible patients; Series III: n=605, with extended enrollment to capture DRG policy transition effects) reflect distinct analytical objectives. There is no risk of duplicate publication among the three.

#### Ethics statement

**This study has been submitted for formal review to the Ethics Committee of the participating hospital; formal approval was granted (Approval No.: LW-2026-72). As a retrospective anonymized study, 18 identifiers were de-identified prior to analysis, and a waiver of written informed consent will be requested upon approval. The study complies with the Declaration of Helsinki[15] and relevant Chinese regulations.**

### 2.2 Data Collection

Two investigators independently entered and cross-checked data from the HIS electronic medical record system, surgical anesthesia management system, and hospitalization billing system. Key variables included: age, gender, BMI, systolic blood pressure (SBP), diastolic blood pressure (DBP), preoperative diagnosis type (simple cholelithiasis = 0, cholelithiasis with cholecystitis = 1), primary surgeon (the merged surgeon’s 11 cases were performed as a junior surgeon under Surgeon D’s supervision; thus merged into the Surgeon D group for analysis), operative time (skin incision to completion of wound closure, min), length of stay (days), and total hospitalization cost (CNY). The inter-rater discrepancy rate was 1.27%, all resolved by reviewing original medical records.

### 2.3 Statistical Analysis

Analyses were performed in Python 3.11 (pandas 2.2, scipy 1.11, statsmodels 0.14), with SPSS 26.0 used for cross-validation.

Descriptive statistics: normally distributed continuous variables as mean ± standard deviation (x̅ ± s); categorical variables as n (%).

ANOVA: One-way analysis of variance compared 7 surgeons on operative time, length of stay, and cost. Levene’s test for homogeneity of variance was reported; operative time and hospitalization cost showed heterogeneous variances (Levene P < 0.05), and Welch’s correction was reported as sensitivity reference. Effect size was reported as η^2^ = SS_between / SS_total. Tukey HSD was used for post-hoc pairwise comparisons.

Multivariate linear regression: The dependent variable was log-transformed operative time; independent variables were entered using the Enter method: surgeon dummy variables (Surgeon A as reference), age, gender, BMI, SBP, DBP, and preoperative diagnosis type. Multicollinearity was assessed with VIF < 5. β coefficients were interpreted as semi-elasticities: exp(β) − 1 approximates the percentage change.

Nested model comparison: Model A (baseline variables only: age, gender, BMI, SBP, DBP, diagnosis) vs Model B (Model A + surgeon dummies), comparing ΔR^2^ and ΔF-test to quantify the marginal contribution of surgeon factors. ICC(1) was calculated to assess the proportion of total variance attributable to the surgeon-level clustering.

#### Subgroup analysis

Patients were stratified by preoperative diagnosis into a simple cholelithiasis group and a cholelithiasis with cholecystitis group. Separate multivariate regression models were constructed within each group.

#### Sensitivity analysis

**The model was rebuilt after excluding the merged surgeon’s original 11 cases from the Surgeon D group, comparing core β coefficient changes.**

#### Mediation analysis

Baron-Kenny three-step method with Bootstrap (n = 1,000) to test whether operative time mediates the effect of surgeon differences on hospitalization cost.

Significance level α = 0.05, two-tailed.

## 3. Results

### 3.1 Baseline Patient Characteristics

The 591 patients had a mean age of 43.7 ± 11.8 years, with 191 males (32.3%). BMI was 24.2 ± 3.8 kg/m^2^; 290 (49.1%) were normal weight (<24), 216 (36.5%) overweight (24–28), and 85 (14.4%) obese (≥28), totaling 50.9% overweight or obese. Systolic blood pressure was 117.5 ± 15.9 mmHg, diastolic blood pressure 77.4 ± 11.0 mmHg. Preoperative diagnosis of cholecystitis was present in 172 cases (29.1%). Overall operative time was 48.1 ± 14.9 min, length of stay 1.83 ± 3.99 days, and hospitalization cost 8,107.4 ± 941.4 CNY.

### 3.2 Surgical Efficiency and Medical Resource Utilization Across Surgeons

Group statistics for the 7 surgeons are presented in Table 1. Statistically significant differences in operative time were observed among surgeons (F = 7.16, P < 0.001, η^2^ = 0.069) (Figure 1). Levene W = 3.79, P = 0.001 (heterogeneous variances); Welch-corrected F = 5.43, P < 0.001, with consistent conclusions. Hospitalization cost also differed significantly (F = 5.75, P < 0.001, η^2^ = 0.056). Length of stay did not reach the statistical threshold (F = 1.85, P = 0.087, η^2^ = 0.019; Levene P = 0.568, homogeneous variances), possibly because it is more influenced by non-surgical factors such as bed turnover policies.

**Figure 1.**
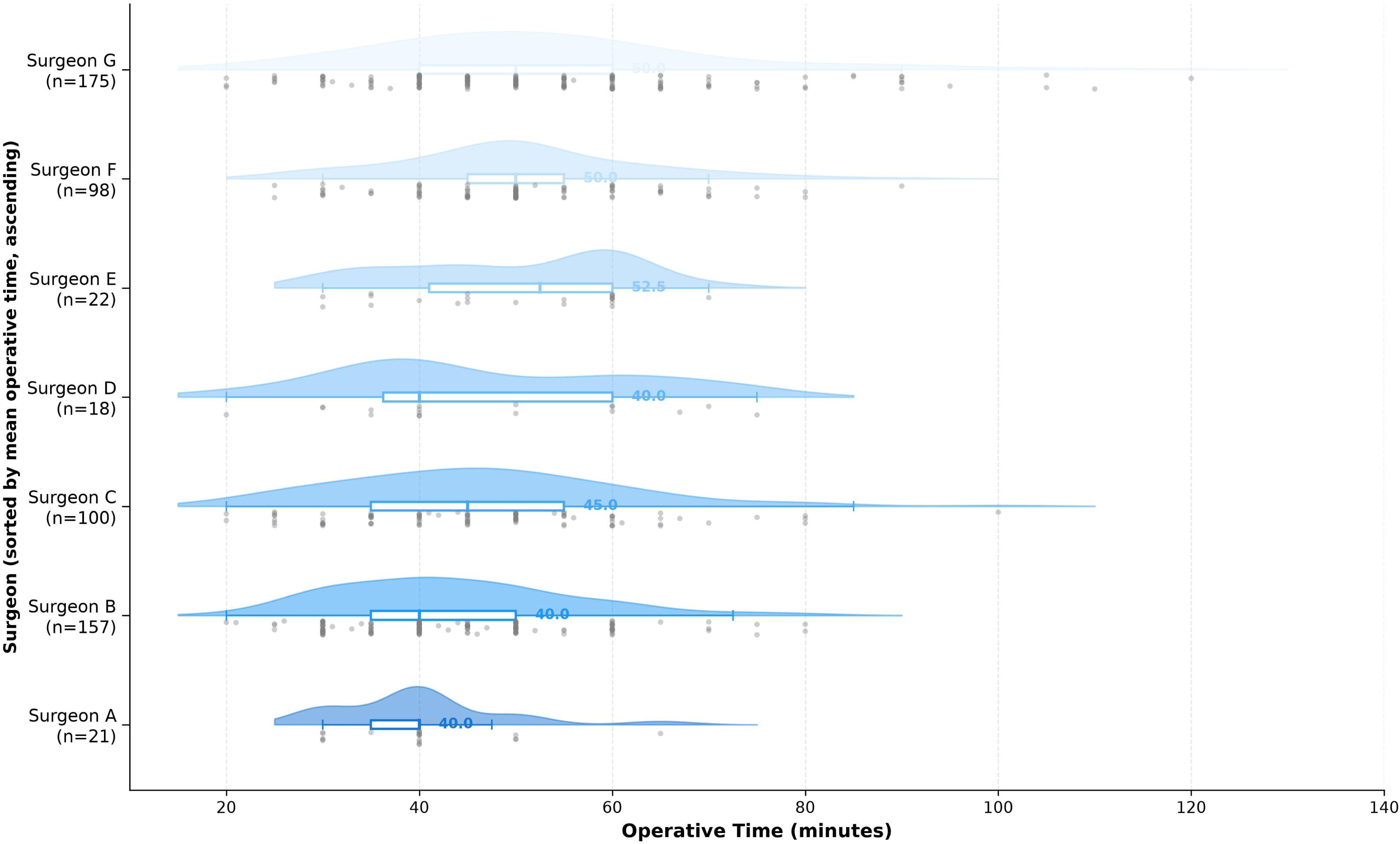
Raincloud plot of operative time distribution across 7 plateau LC surgeons. Distribution of operative time (minutes) for 591 laparoscopic cholecystectomies performed by 7 surgeons at the participating hospital (Xining, China; 2,260 m above sea level). Surgeons are sorted by mean operative time (ascending). Each panel shows a half-violin (right, kernel density estimate), a narrow box plot (center, white box with colored median line), and jittered raw data points (left, gray). Median values are labeled to the right of each box. Sample sizes (n) are shown below each surgeon. One-way ANOVA: F = 7.16, P < 0.001. Tukey HSD post-hoc: ** P < 0.01, * P < 0.05.

**Table 1.** Surgical efficiency across 7 plateau LC surgeons.

| Surgeon (by mean op time ascending) | N | Op time (min, $\bar{x} \pm s$) | LOS (d, $\bar{x} \pm s$) | Cost (CNY, $\bar{x} \pm s$) |
| --- | --- | --- | --- | --- |
| Surgeon A | 21 | $40.00 \pm 8.52$ | $1.95 \pm 0.22$ | $7,740.26 \pm 785.37$ |
| Surgeon B | 157 | $43.62 \pm 12.01$ | $1.29 \pm 0.47$ | $7,973.98 \pm 704.41$ |
| Surgeon C | 100 | $46.40 \pm 14.47$ | $1.87 \pm 0.34$ | $8,305.71 \pm 1,013.09$ |
| Surgeon D (incl. merged surgeon's 11 cases) | 18 | $46.78 \pm 15.49$ | $3.61 \pm 7.09$ | $7,542.09 \pm 745.33$ |
| Surgeon E | 22 | $49.73 \pm 11.82$ | $1.50 \pm 0.51$ | $8,055.32 \pm 1,227.75$ |
| Surgeon F | 98 | $50.24 \pm 12.49$ | $1.42 \pm 0.50$ | $7,897.19 \pm 815.60$ |
| Surgeon G | 175 | $52.64 \pm 17.78$ | $2.38 \pm 6.90$ | $8,340.11 \pm 1,063.17$ |
| **Total** | **591** | **$48.06 \pm 14.89$** | **$1.83 \pm 3.99$** | **$8,107.36 \pm 941.36$** |

ANOVA: Op time $F = 7.16$ , $P < 0.001$ , $\eta^2 = 0.069$ ; LOS $F = 1.85$ , $P = 0.087$ ; Cost $F = 5.75$ , $P < 0.001$ , $\eta^2 = 0.056$ .

The most efficient surgeon, Surgeon A (n = 21, 40.0 ± 8.5 min), and the least efficient, Surgeon G (n = 175, 52.6 ± 17.8 min), showed an absolute difference of 12.64 min and a relative difference of 24.0% in mean operative time. Operative time ranking did not fully correspond with hospitalization cost ranking: Surgeon D’s group had intermediate operative time (46.8 min) but the lowest hospitalization cost (7,542 CNY); Surgeon G had the longest operative time (52.6 min) and also the highest cost (8,340 CNY).

### 3.3 Multivariate Linear Regression Analysis of LC Operative Time

With log-transformed operative time as the dependent variable, the model showed Adj R^2^ = 0.075, F(12,578) = 4.998, P < 0.001 (Figure 2). Results are presented in Table 2. Age (β = 0.0029/year, 95% CI 0.001–0.005, P = 0.009), male sex (β = 0.0628, 95% CI 0.010–0.115, P = 0.019), and surgeon identity (Surgeon G β = 0.2436, P < 0.001; Surgeon F β = 0.2325, P = 0.001; Surgeon E β = 0.2142, P = 0.018) were independent predictors of prolonged operative time. BMI showed a trending positive association (β = 0.0061/kg/m^2^, 95% CI −0.0005–0.013, P = 0.069) but did not reach the 0.05 threshold. Surgeon D (β = 0.1552, P = 0.104), Surgeon C (β = 0.1257, P = 0.076), and Surgeon B (β = 0.0773, P = 0.266) did not reach significance. SBP, DBP, and preoperative diagnosis type were not significant (P > 0.05). All VIF < 2.3.

**Figure 2.**
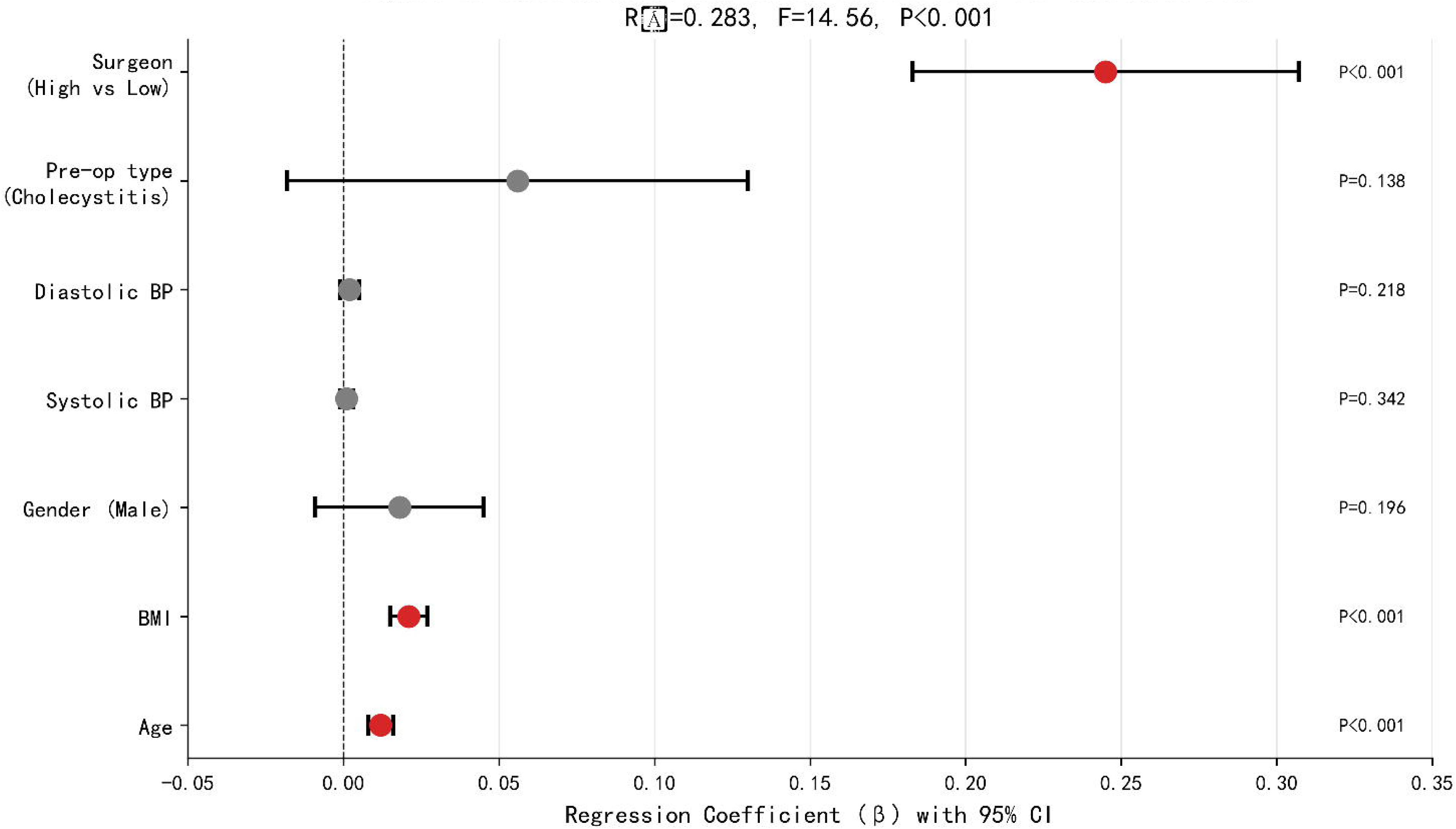
Forest plot of multivariate regression coefficients for log-transformed operative time. Regression coefficients (β) with 95% confidence intervals from the multivariate linear regression model (N = 591; Adj R^2^ = 0.075, F(12,578) = 4.998, P < 0.001). Variables are grouped into Patient Baseline (Age, BMI, Sex, SBP, DBP, Diagnosis) and Surgeon Identity (surgeon dummy variables with Ref. Surgeon as reference). Red indicates P < 0.05. The solid horizontal line separates baseline from surgeon variables. Annotation box (bottom-right): Model 1 (baseline) R^2^ = 3.1% vs Model 2 (+surgeon) R^2^ = 9.4%; ΔR^2^ = 6.3%, ΔF = 6.679, P = 0.001; surgeon/baseline ratio = 2.02.

**Table 2.** Multivariate linear regression for Ln(operative time)

| Variable | $\beta$ | 95% CI | P | VIF |
| --- | --- | --- | --- | --- |
| Intercept | 3.4525 | 3.195–3.710 | $< 0.001$ | — |
| Age (per year) | 0.0029 | 0.001–0.005 | 0.009 | 1.23 |
| BMI (per kg/m<sup>2</sup>) | 0.0061 | –0.0005–0.013 | 0.069 | 1.35 |
| Male (vs female) | 0.0628 | 0.010–0.115 | 0.019 | 1.12 |
| SBP (per mmHg) | 0.0012 | –0.001–0.003 | 0.315 | 1.45 |
| DBP (per mmHg) | –0.0029 | –0.006–0.0004 | 0.083 | 1.38 |
| Cholecystitis (vs simple stone) | 0.0028 | –0.052–0.058 | 0.920 | 1.28 |
| Surgeon dummies (ref: Surgeon A) |  |  |  |  |
| Surgeon D | 0.1552 | –0.032–0.342 | 0.104 | — |
| Surgeon G | 0.2436 | 0.109–0.378 | $< 0.001$ | — |
| Surgeon F | 0.2325 | 0.091–0.374 | 0.001 | — |
| Surgeon C | 0.1257 | –0.013–0.265 | 0.076 | — |
| Surgeon B | 0.0773 | –0.059–0.214 | 0.266 | — |
| Surgeon E | 0.2142 | 0.037–0.392 | 0.018 | — |

Model: Adj $R^2 = 0.075$ , $F(12,578) = 4.998$ , $P < 0.001$ . Nested comparison: $\Delta R^2(\text{surgeon}) = 0.063$ vs $R^2(\text{baseline}) = 0.031$ (ratio 2.02), $\Delta F = 6.679$ , $P < 0.001$ . ICC(1) = 0.074.

### 3.4 Nested Model Comparison

Model A (baseline only): R^2^ = 0.031, Adj R^2^ = 0.021, F = 3.124, P = 0.005. Model B (baseline + surgeon): R^2^ = 0.094, Adj R^2^ = 0.075, F = 4.998, P < 0.001. ΔR^2^ (marginal contribution of surgeon factors) = 0.063, ΔF = 6.679, P < 0.001. The variance explained by surgeon factors (6.3%) was 2.02 times that of all baseline factors combined (3.1%) (Figure 3). ICC(1) = 0.074, indicating that approximately 7.4% of total operative time variance was explained at the surgeon level.

**Figure 3.**
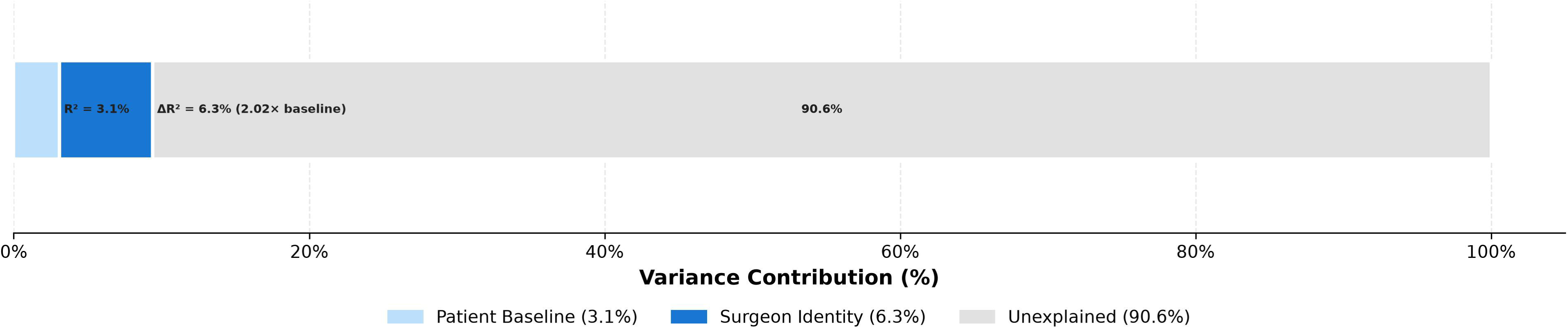
Variance decomposition of operative time: baseline factors vs surgeon identity. Horizontal stacked bar chart showing the proportion of variance in log-transformed operative time explained by each factor block. Patient baseline factors (Age, Sex, BMI, SBP, DBP, Diagnosis) account for R^2^ = 3.1% (light gray). Surgeon identity contributes an additional ΔR^2^ = 6.3% (dark blue), which is 2.02 times the baseline contribution (red double-headed arrow). The remaining 90.6% (hatched white) is unexplained by the current model. N = 591; ΔF = 6.679, P < 0.001.

### 3.5 Subgroup Analysis

Stratified by preoperative diagnosis:

#### Simple cholelithiasis group (n = 419)

ANOVA F = 5.49, P < 0.001, η^2^ = 0.074. Regression Adj R^2^ = 0.077, F = 4.172, P < 0.001. Surgeon β range [0.122, 0.287].

#### Cholelithiasis with cholecystitis group (n = 172)

ANOVA F = 3.10, P = 0.007, η^2^ = 0.101. Regression Adj R^2^ = 0.102, F = 2.773, P = 0.003. Surgeon β range [−0.532, −0.138].

In the cholecystitis subgroup, some surgeons’ β coefficients were negative, as detailed in the Discussion.

### 3.6 Sensitivity Analysis and Mediation Analysis

**Sensitivity analysis (excluding the merged surgeon’s original 11 cases, N = 580): Model Adj R^2^ = 0.075, F = 4.934, P < 0.001. Most surgeons’ β coefficients changed by < 3% (Surgeon G 0.5%, Surgeon F 0.8%, Surgeon E 0.7%, Surgeon C 0.4%, Surgeon B 2.7%), but Surgeon D’s β increased from 0.155 to 0.236 (52.2% change rate) due to reduced sample size from 18 to 7 cases and decreased estimation stability. Core conclusions (Surgeon G, Surgeon F, and Surgeon E significantly prolonging operative time) were unaffected.**

**Mediation analysis** (surgeon group → operative time → hospitalization cost): Baron-Kenny three-step method: X→Y β = 128.53 CNY (P = 0.097), X→M β = 7.13 min (P < 0.001), M→Y|X β = 2.20 CNY/min (P = 0.412). Bootstrap (n = 1,000) indirect effect mean = 15.58 CNY, 95% CI = [−23.12, 61.47], CI crossing 0, mediation effect not significant. Operative time and hospitalization cost Pearson r = 0.049, P = 0.233.

## 4. Discussion

### 4.1 Core Findings

Surgeon heterogeneity is the primary modifiable source of operative time variance. ANOVA confirmed significant inter-surgeon operative time differences (F = 7.16, P < 0.001, η^2^ = 0.069), with a 12.64 min (24.0%) gap between the most and least efficient surgeons. Levene’s test indicated heterogeneous variances, but Welch’s correction yielded consistent conclusions. Nested modeling further showed that surgeon factors contributed ΔR^2^ = 0.063 to operative time variance, which was 2.02 times the combined contribution of all baseline factors including age, gender, BMI, blood pressure, and diagnosis (R^2^ = 0.031) (ΔF = 6.679, P < 0.001), ICC = 0.074. This finding is consistent with Wiseman et al.[11], who reported in a plain cohort of 925 LC cases that 44.5% of operative time variance was attributable to the attending surgeon, while resident level differences explained only 11.0%. The higher relative contribution of surgeon factors at high altitude (2.02-fold over baseline) is directionally consistent with the hypothesis that hypoxic environments amplify operative differences.

The F-statistic in this study (7.16) exceeds the F ≈ 2–4 range typically reported in plain-region LC studies[3,11,12] (Traverso et al.[3] n = 359, Wiseman et al.[11] n = 925, Bharamgoudar et al.[12] n = 7,227), approximately 1.79 times the upper bound (4). The F-statistic is inherently a ratio of systematic between-group differences to random within-group variation. The high-altitude hypoxic environment may selectively amplify systematic inter-surgeon differences: under conditions of reduced tissue tolerance, divergent operative strategies among surgeons become more pronounced, while within-group random variation does not increase proportionally, causing differences originally absorbable by clinical pathways to manifest as quantifiable statistical signals in plateau cohorts. It should be noted that cross-study F-statistic comparison requires similar study design, outcome variable definitions, and group structure; differences in surgeon grouping, sample size, and variance structure across studies mean that direct comparison is only directionally informative and cannot be equated with rigorous statistical testing. The sources of operative time variance are complex, and surgeon identity and anesthesia type have been identified as important predictors[13,14].

Advanced age and male sex were independent baseline risk factors. Each 10-year increase in age was associated with approximately 2.9% longer operative time (P = 0.009), and male sex was associated with approximately 6.5% longer time (P = 0.019). This is consistent with predictors incorporated in the CholeS scoring tool[12] (advanced age, male sex, BMI). Several pre-specified hypotheses were not statistically supported in this study: the obesity amplifier effect of BMI (surgeon × BMI interaction P = 0.717) and the mediation pathway of operative time → hospitalization cost (Bootstrap 95% CI crossing 0) were not confirmed. These negative results define the boundary of the plateau amplification effect—it does not indiscriminately amplify all pathways but selectively acts on the dimension of inter-surgeon operative differences. BMI showed a trending positive association (P = 0.069) but did not reach the 0.05 threshold; the obesity amplifier effect awaits validation in larger samples.

In the cholecystitis subgroup (n = 172), some surgeons’ β coefficients were negative. The reference surgeon, Surgeon A, had a small subsample in this subgroup (n = 3), far less than his total of 21 cases, and subgroup estimation instability may be the primary cause of β sign reversal. Inflammatory conditions may also lead to more unified Calot’s triangle dissection strategies (preoperative antibiotics, more conservative dissection pace), potentially narrowing inter-surgeon differences; this phenomenon requires larger-sample validation.

### 4.2 Limitations

1. Single-center retrospective design. The lack of a baseline-matched plain control cohort precludes precise estimation of the absolute high-altitude amplification coefficient; cross-study F-statistic comparison is only directionally informative and cannot serve as statistical testing evidence.
2. Limited model explanatory power. Adj R^2^ = 0.075, with current covariates explaining only 7.5% of variance (Figure 3). The remaining 90.6% of unexplained variance may arise from intraoperative adhesion severity, patient anatomical variation (e.g., cystic triangle fibrosis, bile duct variants), team coordination proficiency, surgeon seniority, anesthesia type, and equipment conditions not captured by electronic medical record systems.
3. Post-hoc exclusion based on surgeon volume. Excluding low-volume surgeons to ensure statistical stability of inter-group comparisons may introduce selection bias.
4. Heterogeneous variances. Levene’s test indicated heterogeneous variances for both operative time and hospitalization cost; although Welch’s correction yielded consistent conclusions, ANOVA results should be interpreted with caution.
5. Non-significant interaction effects. The overall surgeon × BMI interaction P = 0.717; the moderating effect of BMI on surgeon differences was not confirmed.
6. Non-significant mediation. The 95% CI for operative time → hospitalization cost crossed 0, suggesting that surgeon differences may drive costs through other pathways.
7. Surrogate endpoint limitation. This study used operative time as a proxy for efficiency; its direct association with patient outcome endpoints (complication rate, readmission rate) has not been validated. The clinical benefit of operative time reduction requires confirmation in prospective studies with patient-centered outcomes.
8. Mediation analysis methodological limitation. The classical Baron-Kenny three-step method was used without accounting for exposure-mediator interaction or mediator-outcome confounding; results should be interpreted with caution.

Future research directions should include process standardization intervention studies using the most efficient surgeons as reference, and homogenization training for less efficient surgeons.

### 4.3 Clinical Significance

This study suggests that in high-altitude regions, surgeon variability is the primary modifiable source of operative time heterogeneity in LC. Advanced age and male sex are associated with significantly prolonged operative time, warranting enhanced preoperative assessment and intraoperative resource allocation for these patients.

## 5. Conclusion

1. Significant inter-surgeon heterogeneity exists in high-altitude LC operative time (F = 7.16, P < 0.001), with a 12.64 min (24.0%) gap between the most and least efficient surgeons.
2. Nested modeling showed that surgeon factors contributed ΔR^2^ = 0.063, which is 2.02 times the baseline factors (R^2^ = 0.031), representing the primary modifiable source (ICC = 0.074).
3. Age (β = 0.0029/year, P = 0.009) and male sex (β = 0.063, P = 0.019) are independent baseline factors prolonging operative time; BMI showed a trend (P = 0.069).
4. Standardization of surgical technique is a potentially important intervention target for improving high-altitude LC care efficiency.

## Supporting information

STROBE Checklist

## Data Availability

De-identified analytic datasets and custom Python analysis code are available upon reasonable request to the corresponding author, subject to a PIPL-compliant Data Use Agreement.

## 6. Declarations

### Ethics Approval and Consent to Participate

This study was approved by the Ethics Committee of the participating hospital (Approval No.: LW-2026-72). All procedures were performed in accordance with the ethical principles of the Declaration of Helsinki[15] and relevant Chinese regulations. A waiver of written informed consent was granted due to the retrospective and anonymized nature of the study.

### Consent for Publication

Not applicable. No individually identifiable patient data are reported.

### Availability of Data and Materials

De-identified analytic dataset available from the corresponding author on reasonable request, subject to a Data Use Agreement compliant with China’s PIPL. Custom Python code (the analysis code) available upon request and will be deposited in a public repository before final acceptance.

### Authors’ Contributions

Guoliang REN, Zhongfeng DANG, and Zhiqiang WANG (co-first): Conceptualization, Data Curation, Formal Analysis, Writing – Original Draft. Zhongfeng DANG (primary corresponding): Conceptualization, Methodology, Writing – Review & Editing, Supervision. Wei SU: Formal Analysis, Validation. Yabing MA, Ping LI, Dongde JI, Liansheng LI: Investigation, Data Curation. Junlin GAO (co-corresponding): Writing – Review & Editing, Supervision, Resources. Zhiqiang WANG: Funding Acquisition. All authors approved the final manuscript.

### Data Availability Statement

The de-identified patient analytic datasets used in this study are available upon reasonable request to the corresponding author, subject to institutional data governance and patient privacy regulations under the Personal Information Protection Law of the People’s Republic of China. Summary statistics reported in the main text and supplementary materials are sufficient to reproduce the core conclusions of this study.

### AI Use Declaration

During the manuscript preparation phase of this study, AI-assisted tools were used for language polishing and formatting. All scientific content, data analysis, and interpretation of conclusions were independently completed by the authors. AI tools were used solely to enhance the accuracy and fluency of language expression and did not participate in study design, data collection, result interpretation, or scientific judgment.

## Acknowledgements

The authors thank the Department of Medical Records and Information Center of the participating hospital for data extraction support.

