## Supplementary material for "Operative Time Heterogeneity in Laparoscopic Cholecystectomy at High Altitude: Surgeon Variability as a Modifiable Factor under Hypoxic Stress": STROBE Checklist

### STROBE Checklist — Paper 1

Reporting Guideline: STROBE (Strengthening the Reporting of Observational Studies in Epidemiology) — Observational Cohort Study

Checklist Version: STROBE 2007 (updated 2014)

Last Updated: 2026-08-16

---

#### Checklist

| Item | Recommendation | Page/Section | Reported |
|---|---|---|---|
| **Title and abstract** | | | |
| 1 | (a) Indicate study design with a commonly used term in the title or abstract | Abstract (Methods) | ✅ |
| 1 | (b) Provide in the abstract an informative and balanced summary of what was done and what was found | Abstract | ✅ |
| **Introduction** | | | |
| 2 | Explain the scientific background and rationale for the investigation being reported | Introduction | ✅ |
| 3 | State specific objectives, including any prespecified hypotheses | Introduction (last paragraph) | ✅ |
| **Methods** | | | |
| 4 | Present key elements of study design early in the paper | Methods (Study Design) | ✅ |
| 5 | Describe the setting, locations, and relevant dates, including periods of recruitment, exposure, follow-up, and data collection | Methods (Setting) | ✅ |
| 6 | (a) Give the eligibility criteria, and the sources and methods of selection of participants | Methods (Participants) | ✅ |
| 7 | Clearly define all outcomes, exposures, predictors, potential confounders, and effect modifiers | Methods (Variables) | ✅ |
| 8 | For each variable of interest, give sources of | Methods (Data Sources) | ✅ |

| | | | |
|---|---|---|---|
| | data and details of methods of assessment | | |
| 9 | Describe any efforts to address potential sources of bias | Methods (Bias) | ✅ |
| 10 | Explain how the study size was arrived at | Methods (Sample Size) | ✅ |
| 11 | Explain how quantitative variables were handled in the analyses. If relevant, describe which groupings were chosen and why | Methods (Statistical Analysis) | ✅ |
| 12 | (a) Describe all statistical methods, including those used to control for confounding | Methods (Statistical Analysis) | ✅ |
| 12 | (b) Describe any methods used to examine subgroups and interactions | Methods (Nested Model Comparison) | ✅ |
| 12 | (c) Explain how missing data were addressed | Methods (Missing Data) | ✅ |
| 12 | (d) If applicable, describe analytical methods taking account of sampling strategy | N/A (single-center) | N/A |
| 12 | (e) Describe any sensitivity analyses | Methods (Sensitivity Analysis) | ✅ |
| **Results** | | | |
| 13 | (a) Report numbers of individuals at each stage of study—eg numbers potentially eligible, examined for eligibility, confirmed eligible, recruited into the study, completing follow-up, and analysed | Results (Participants) | ✅ |
| 13 | (b) Give reasons for non-participation at each stage | Results (Participants) | ✅ |
| 13 | (c) Consider use of a flow diagram | N/A (retrospective) | N/A |
| 14 | (a) Give characteristics of study participants (eg demographic, clinical, social) and information on exposures and potential confounders | Results (Table 1) | ✅ |
| 14 | (b) Indicate number of participants with missing data for each variable of interest | Results (Table 1 footnote) | ✅ |
| 15 | Report numbers of outcome events or summary measures over time | Results (Outcome Data) | ✅ |
| 16 | (a) Give unadjusted estimates and, if applicable, confounder-adjusted estimates and their precision (eg 95% confidence interval) | Results (Table 2) | ✅ |

| | | | |
|---|---|---|---|
| 16 | (b) Report category boundaries when continuous variables were categorized | Results (Table 2 footnote) | ✅ |
| 16 | (c) If relevant, consider translating estimates of relative risk into absolute risk for a meaningful time period | N/A | N/A |
| 17 | Report other analyses done—eg analyses of subgroups and interactions, and sensitivity analyses | Results (Nested Model, ICC) | ✅ |
| **Discussion** | | | |
| 18 | Summarise key results with reference to study objectives | Discussion (Key Findings) | ✅ |
| 19 | Give a cautious overall interpretation of results considering objectives, limitations, multiplicity of analyses, results from similar studies, and other relevant evidence | Discussion (Interpretation) | ✅ |
| 20 | Discuss limitations of the study, taking into account sources of potential bias or imprecision | Discussion (Limitations) | ✅ |
| 21 | Give a cautious overall interpretation of results | Discussion (Generalizability) | ✅ |
| **Other information** | | | |
| 22 | Give the source of funding and the role of the funders | Funding (YNZXKT2026009) | ✅ |

---

### Notes

- This is an observational retrospective cohort study; STROBE is the appropriate reporting guideline.
- The study was conducted at Qinghai Red Cross Hospital (altitude 2,260 m) from 2020 to 2023.
- Total participants: 591 elective LC cases by 7 surgeons.
- Ethics Approval: LW-2026-72 (Qinghai Red Cross Hospital Medical Ethics Committee, 2026-08-13).

---

Checklist completed on 2026-08-16 by Zhongfeng DANG.
